# Antibody Attributes that Predict the Neutralization and Effector Function of Polyclonal Responses to SARS-CoV-2

**DOI:** 10.1101/2021.08.06.21261710

**Authors:** Harini Natarajan, Shiwei Xu, Andrew R. Crowley, Savannah E. Butler, Joshua A. Weiner, Evan M. Bloch, Kirsten Littlefield, Sarah E. Benner, Ruchee Shrestha, Olivia Ajayi, Wendy Wieland-Alter, David Sullivan, Shmuel Shoham, Thomas C. Quinn, Arturo Casadevall, Andrew Pekosz, Andrew D. Redd, Aaron A.R. Tobian, Ruth I. Connor, Peter F. Wright, Margaret E. Ackerman

**Affiliations:** Department of Microbiology and Immunology, Geisel School of Medicine at Dartmouth, Dartmouth College, Hanover, NH, USA; Program in Quantitative Biological Sciences, Dartmouth College, Hanover, NH, USA; Thayer School of Engineering, Dartmouth College, Hanover, NH, USA; Department of Pathology, Johns Hopkins School of Medicine, Baltimore, MD, USA; W. Harry Feinstone Department of Molecular Microbiology and Immunology, Johns Hopkins Bloomberg School of Public Health, Baltimore, MD, USA; Department of Pediatrics, Geisel School of Medicine at Dartmouth, Dartmouth-Hitchcock Medical Center, Lebanon, NH, USA; Department of Medicine, Division of Infectious Diseases, Johns Hopkins School of Medicine, Baltimore, MD, USA; Division of Intramural Research, National Institute of Allergy and Infectious Diseases, National Institutes of Health, Bethesda, MD, USA

**Author notes:** Corresponding Author Margaret E. Ackerman 14 Engineering Drive, Hanover, NH 03755, (ph). Contributed equally (Harini Natarajan, Shiwei Xu).

## Abstract

While antibodies provide significant protection from SARS-CoV-2 infection and disease sequelae, the specific attributes of the humoral response that contribute to immunity are incompletely defined. In this study, we employ machine learning to relate characteristics of the polyclonal antibody response raised by natural infection to diverse antibody effector functions and neutralization potency with the goal of generating both accurate predictions of each activity based on antibody response profiles as well as insights into antibody mechanisms of action. To this end, antibody-mediated phagocytosis, cytotoxicity, complement deposition, and neutralization were accurately predicted from biophysical antibody profiles in both discovery and validation cohorts. These predictive models identified SARS-CoV-2-specific IgM as a key predictor of neutralization activity whose mechanistic relevance was supported experimentally by depletion. Validated models of how different aspects of the humoral response relate to antiviral antibody activities suggest desirable attributes to recapitulate by vaccination or other antibody-based interventions.

## Introduction

The SARS-CoV-2 pandemic has resulted in over 127 million cases, 2.7 million deaths, and unprecedented social, economic, and educational impact despite interventions that have included quarantines, shutdowns, social distancing, and masking requirements. However, the pandemic has also led to international collaborations working toward understanding the disease and developing novel therapeutics and vaccines. To date, these efforts have resulted in several novel therapies and several vaccines approved for widespread deployment under emergency use authorization (EUA)^1^.

The success of these vaccines is thought to result in no small part to the potent antiviral activities of the antibodies they induce. While reinfections have been documented^2,3^, seropositivity and levels of neutralizing antibody are associated with highly reduced rates of re-infection^4-6^, and passive transfer of plasma from convalescent donors has shown therapeutic efficacy in some studies^7-14^ but not others^15-18^. The inconsistent results with convalescent plasma studies suggest that the variables that contribute to passive antibody efficacy in polyclonal preparations are not completely understood. Additionally, built on strong preclinical data showing the ability of antibodies to prevent infection^5^, monoclonal antibody therapies have been developed, including combination products^19,20^. Each of the three vaccines currently under emergency use in the United States induces neutralizing antibodies, often to levels exceeding those detected following natural infection^21-23^.

However, whether elicited by vaccination or infection, antibody responses between individuals are highly variable^24-27^, both in titer and in composition. This variability suggests that monoclonal antibody and convalescent plasma therapy, as well as vaccine design, can be improved by determining the factors that contribute to a functionally protective antibody response. Beyond neutralization, which has been established as a correlate of protection in diverse studies^20,28-30^, evidence has accrued supporting both protective and pathogenic roles of antibody effector functions in infection resistance and disease severity. These functions include activities mediated by both soluble factors and diverse innate immune effector cell types. For example, initiation of the complement cascade can result in direct viral or infected cell lysis^31^, or modification of other activities including neutralization^31,32^. Similarly, antibodies can induce phagocytosis, drive release of cytotoxic factors such as perforin and granzyme B, or secretion of inflammatory mediators such as a cytokines and reactive oxygen species^32,33^. In studies of SARS-CoV-2, extra-neutralizing functions have been shown to play an important role in antiviral activity of antibodies^34-40^. The importance of these functions has been defined *in vivo* in animal models using both using Fc engineering to modulate binding of the Fc domain to Fcγ Receptors (FcγR), and through depletion of effector cells. In contrast, in correlative studies some extra-neutralizing functions have also been linked to disease severity^41,42^. These findings suggest the importance of understanding the role of both neutralization and extra-neutralizing functions in antibody responses to SARS-CoV-2 infection. Given these observations, better understanding of the relationship between the magnitude and character of the humoral immune response and diverse antibody activities may offer key insights to further the development of successful therapeutics and vaccines for SARS-CoV-2.

## Results

### Characterization of antibody responses following SARS-CoV-2 infection

Antibody functions, including neutralization assessed by either an authentic virus assay or a luciferase-based pseudovirus assay, antibody-dependent cell-mediated phagocytosis (ADCP) mediated by monocytes, deposition of the complement cascade component C3b (ADCD), and FcγRIIIa ligation as a proxy for NK cell mediated antibody dependent cellular cytotoxicity (ADCC) induced by antibodies in response to recombinant antigen were previously reported^26^ for a set of convalescent samples collected from a discovery cohort of 126 eligible convalescent plasma donors from the Baltimore/Washington D.C. area (Johns Hopkins Medical Institutions, JHMI)^27^ and serum samples from 15 naïve controls and a validation cohort of 20 convalescent subjects from New Hampshire (Dartmouth-Hitchcock Medical Center, DHMC)^43^ (**Supplemental Table 1**). Biophysical antibody features were defined by a customized multiplexed Fc array assay that characterizes both variable fragment (Fv) and Fc domain attributes across a panel of SARS-CoV-2 antigens, consisting of: nucleocapsid (N) protein, stabilized (S-2P)^44^ and unstabilized trimeric spike protein, spike subdomains including S1 and S2, the receptor binding domain (RBD), and the fusion peptide (FP) from SARS-CoV-2; in addition, the panel included diverse pathogenic, zoonotic, and endemic coronavirus spike proteins and subdomains. Influenza hemagglutinin (HA) and herpes simplex virus glycoprotein E (gE) were evaluated as controls. The Fc domain characteristics evaluated for each antigen specificity included antibody isotype, subclass, and propensity to bind Fc receptors (FcRs) (**Supplemental Table 2**).

To understand how the different facets of the Ab response relate to one another, hierarchical clustering was performed on the biophysical antibody profiles of convalescent plasma donors (JHMI) and compared to the serum profiles of SARS-CoV-2 naïve subjects. Extensive variability in the SARS-CoV-2-specific Ab response magnitude and character was noted (**Figure 1A**). High levels of IgG were observed in many individuals, particularly those who had been hospitalized, while a small number of convalescent donors appeared not to seroconvert despite documented infection via nucleic acid amplification. Similarly, there was considerable variability in the IgA and IgM responses in SARS-CoV-2-convalescent subjects. IgG2, IgG4, and IgD responses were less commonly observed. Distinctions in antibody responses between subjects were apparent among antigen specificities. For example, perhaps due its high homology with endemic CoV, FP responses were isotype switched consistent with an amnestic response, whereas IgM responses to S were reliably observed.

**Figure 1:**
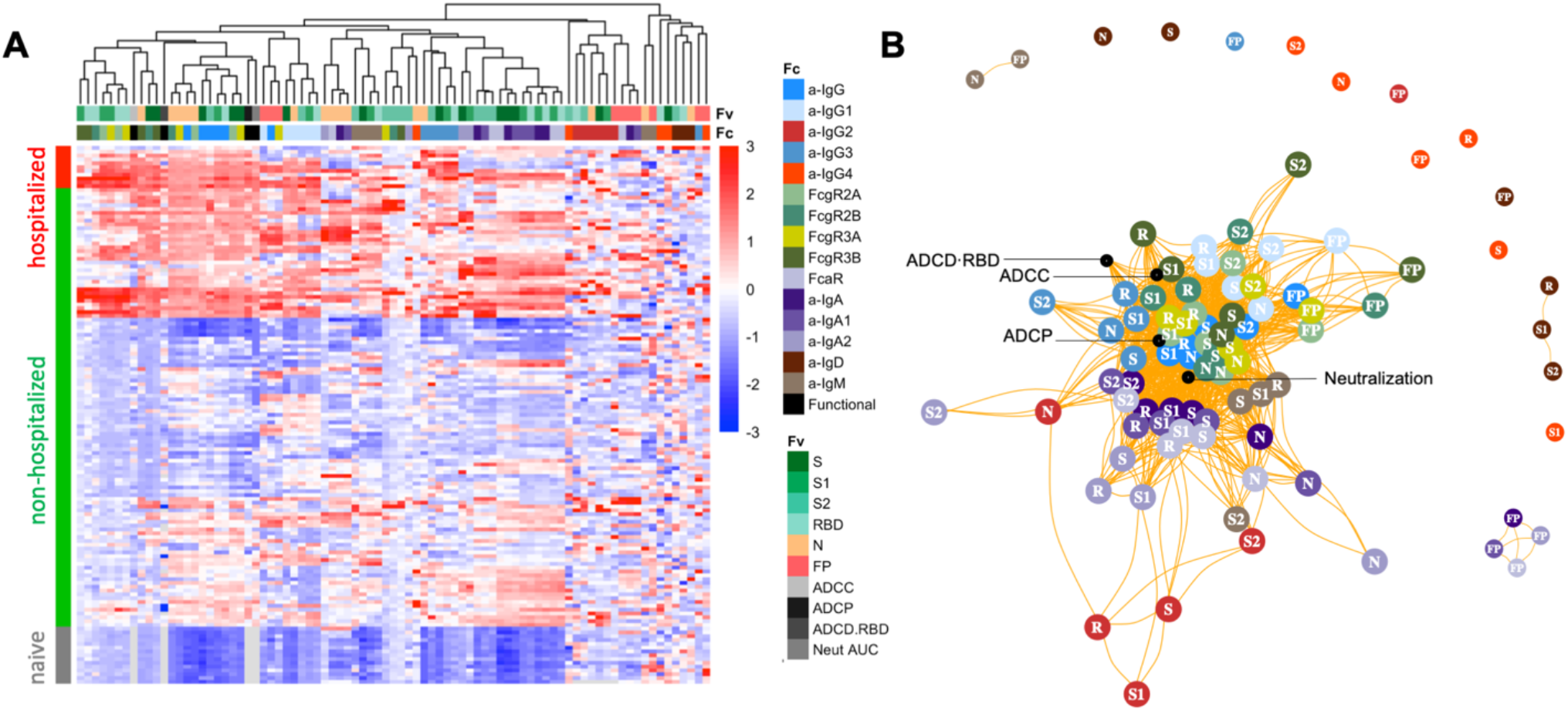
Biophysical and functional antibody responses among convalescent donors. **A**. Heatmap of filtered and hierarchically clustered SARS-CoV-2-specific Fc array features across disease severity and infection status in the JHMI cohort. Each row represents an individual subject, and subjects are grouped by disease status, as indicated by the vertical color bar. Each column represents an Fc array feature; horizontal color bars indicate each function or each Fv-specificity and Fc-characteristic tested. Responses are scaled and centered per feature and the range was truncated +/- 3 SD. Higher responses are indicated in red and lower responses are indicated in blue. Missing data is indicated in light gray. **B**. Weighted network plots of correlative relationships (|r| > 0.5) among antibody functions (black) and CoV-2-specific antibody features. Fc array measurements are colored by Fc characteristic and Fv specificity is indicated in text label (R = RBD).

A weighted network plot depicting Pearson’s correlation coefficients between Fc array features and functional measurements was created to elucidate correlative relationships more directly between aspects of humoral responses (**Figure 1B**). As was apparent in the heatmap (**Figure 1A**), features were often more strongly grouped by Fc domain characteristics than antigen-specificity. Nodes representing antibody effector functions clustered more tightly with RBD-, S1-, S- and N-specific FcγR-binding levels, IgG3, and total IgG responses than with IgG1 responses or those directed at S2 or FP. Though most closely linked to IgG-associated features, neutralization potency appeared as a hub that connected to IgA and IgM responses. Based on both hierarchical clustering and correlation analysis (**Figure 1B**), the ability of antigen-specific antibodies to interact with diverse FcγR was well correlated to multiple antibody effector functions.

### Multivariate modelling methods to predict functional responses

With the dual goals of better understanding the humoral response features that may drive complex antibody functions and enabling robust predictions from surrogate measures, we applied supervised machine learning methods to this (JHMI) dataset, while using the DHMC cohort as validation to determine whether the models could predict activity in a generalized manner. A regularized generalized linear modeling approach trained to utilize Fc Array features to predict each antibody function with minimal mean squared error was selected based on prior success in identifying interpretable factors that contribute to functional activity while avoiding overfitting^45^. Five-fold cross-validation was employed to evaluate generalizability within the JHMI cohort, and comparison to models trained on permuted functional data established model robustness (**Figure 2A**). The cross-validated models trained on diverse data subsets showed similar accuracy (measured by mean squared error) when applied to held out subjects as when used to predict effector function and neutralization activity that was observed in the validation cohort (DHMC). Model quality was also evaluated in terms of the degree of correlation between predicted and observed activity for a representative cross-validation replicate, allowing for better visualization of model performance (**Figure 2B**).

**Figure 2:**
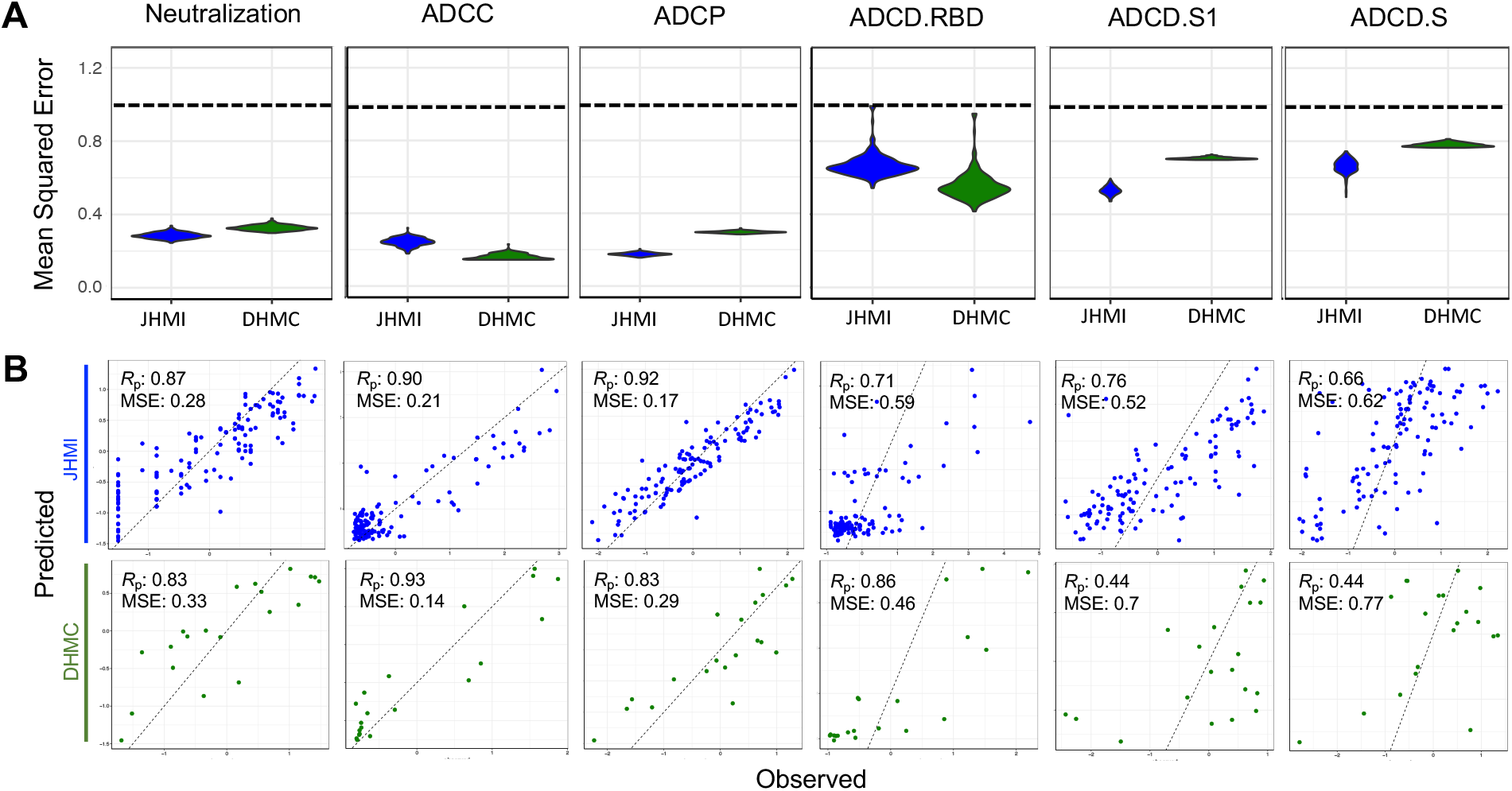
Multivariate linear regression modeling validation in test set. **A**. Comparison of mean-squared error between testing (JHMI) and validation (DHMC) data sets for each functional assay across cross-validation replicates. Dotted line indicates median performance on permuted data in the setting of repeated cross-validation. **B**. Correlation between predicted and observed responses in the discovery (JHMI, blue) and validation (DHMC, green) cohorts. Pearson correlation (Rp) and mean squared error (MSE) are reported in inset. Dotted line indicates x=y.

The model consistently selected a subset of features for each function (**Figure 3A**). The features that appeared with high frequency in repeated modeling were likely to have relatively high coefficients, and inversely, biophysical features with relatively small coefficients were prone to be influenced by the selected sample subset and to be removed by chance across the replicates. Collectively, the frequently contributing features were exclusively related to spike recognition and were primarily driven by IgG and FcγR-binding antibodies.

**Figure 3:**
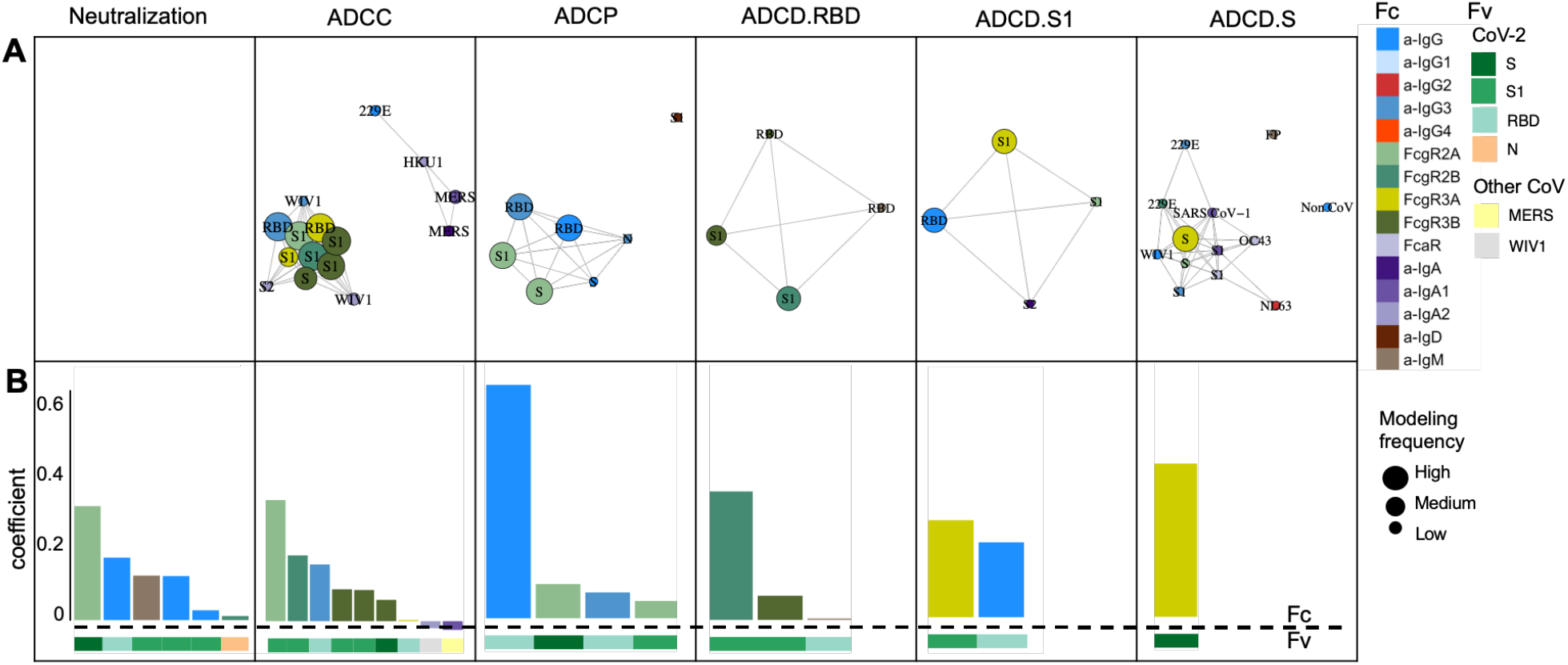
Final predicted biophysical features and contributions in the multivariate linear regression modeling. **A**. Network showing the identity, relative degree of correlation, and frequency with which features contribute to models in the setting of repeated cross-validation. **B**. Coefficients of biophysical features to the final models predictive of each function. Antigen specificity (Fv) and Fc characteristics (Fc) are shown in color bars

To evaluate the magnitudes of feature contributions, a representative model for each function demonstrating the identity and relative coefficients of the contributing features is presented (**Figure 3B**). Again, despite their sparseness compared to the control antigen, endemic CoV, and other epidemic CoV features, these models relied almost exclusively on antibody responses to the SARS-CoV-2 spike. Consistent with the experimental approach evaluating functions elicited specifically against RBD, ADCC and ADCP models depended principally on antibodies specific to RBD or more broadly to S1. In contrast, the lead feature for virus neutralization was recognition of stabilized spike (S-2P). Similarly, complement deposition against whole spike was best predicted by a single feature related to spike trimer recognition. Responses to the S2 domain were not observed to contribute to functional predictions. Intriguingly, IgA responses against other CoV were observed to make inverse contributions to ADCC predictions. While these contributions were of small magnitude, this result suggests the possibility that cross-reactive, potentially S2-specific IgAs may inhibit the activity of S-reactive IgGs, as has been observed in the context of the HIV envelope glycoprotein^46^.

Beyond specificity, distinct antibody Fc characteristics contributed to model predictions. The most frequent Fc characteristic of features contributing to the final model of neutralization potency was the magnitude of IgG response, consistent with neutralization being FcR-independent. In contrast, the most frequent Fc characteristics in modeling ADCC and ADCP were FcγRIII- and FcγRII-binding responses, respectively – the receptors most relevant to each function. Further, despite comprising a relatively small fraction of circulating IgG, but consistent with its enhanced ability to drive effector functions^47,48^, IgG3 antibodies specific to RBD made a substantial contribution to models of both ADCP and ADCC activity, suggesting the potential importance of this subclass. Intriguingly, S1-specific IgM contributed to models of neutralization potency. IgM is typically associated with initial exposures^49^, and our data suggesting the possibility that this feature represents *de novo* rather than recalled cross-reactive lineages that may exhibit superior neutralization activity, as has been observed in the context of influenza responses^50,51^. Overall, while functions were predicted with differing degrees of accuracy, each generalized well to the independent validation cohort and relied upon features with established biological relevance.

### Experimental validation of predictive models of antibody function

Given the somewhat surprising appearance of an IgM feature in predictions of neutralization activity, we sought to evaluate the mechanistic relevance of this isotype in particular. In a select group of individuals with both high IgM and neutralization levels (n=11), IgM was depleted from serum to determine whether the loss of CoV-2-specific IgM resulted in a reduction in neutralization of SARS-CoV-2 (**Supplemental Figure 1**). Both total (not shown) and RBD-specific IgM was depleted (97-fold) (**Figure 4A**). Minimal effects on total (not shown) and RBD-specific IgG (2.0-fold) and IgA (2.3-fold) levels were observed in the IgM-depleted samples. Following IgM depletion, samples showed 1.6-to 73-fold decreases in neutralization titer (**Figure 4B**). Though the magnitude of changes in Ig levels and neutralization before and after depletion varied per donor, only IgM and not IgG or IgA levels showed a statistically significant correlation with neutralization titer in these individuals (**Figure 4C**). This result demonstrates that mechanistically relevant features can be discovered from unbiased data analysis and modeling processes.

**Figure 4:**
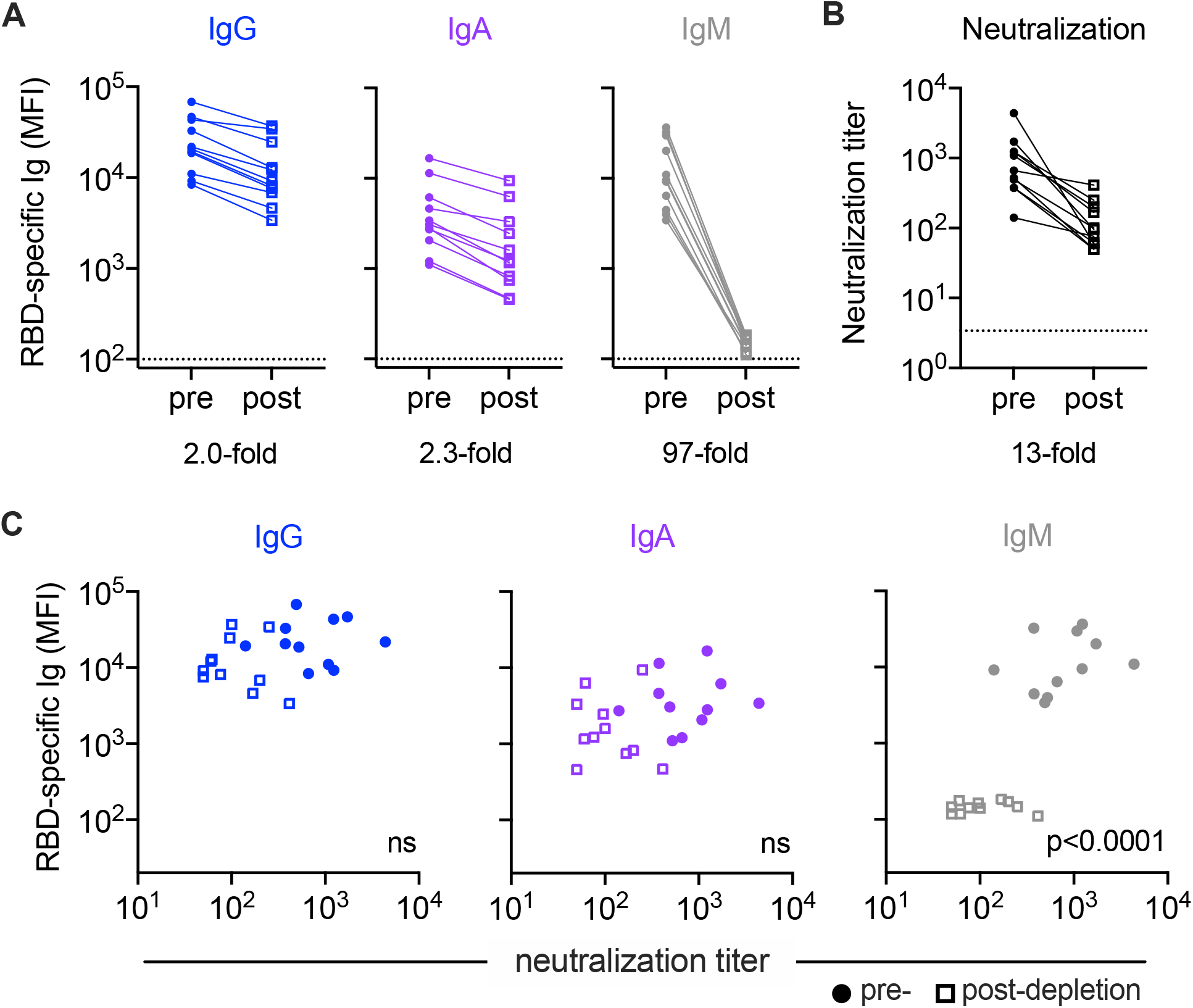
Experimental validation of IgM-mediated neutralization. **A**. Median fluorescent intensity (MFI) levels of RBD-specific IgG, IgA, IgM observed pre- (filled circles) and post-(hollow squares) IgM depletion. Mean fold change in MFI across samples for each isotype is indicated below the figure. **B**. Neutralization titers pre-and post-IgM depletion. **C**. Comparison of RBD-specific Ig levels to neutralization titer. Statistical significance (two-tailed p value) of Spearman correlation coefficients reported in inset.

## Discussion

It is now well established that SARS-CoV-2-specific antibodies can drive varied antiviral functions beyond neutralization^34,43,52^. These responses have been less well characterized, but accumulating evidence suggests their importance to protection from infection and disease. Both ADCC and phagocytosis have been reported to contribute to antibody-mediated antiviral activity against other coronaviruses^53-55^. Collectively, these functions have been suggested to play an important role in defense against SARS-CoV-2; they have been implicated in *in vivo* protection in diverse studies, including passive transfer studies that have demonstrated that effector functions play a role in the antiviral activity of monoclonal antibodies and correlates of protection analysis carried out on vaccine candidates^34-40^. Fc engineering approaches that both knocked out or enhanced antibody effector functions and studies of the depletion of effector cells in the context of diverse antibodies have provided convincing evidence of the mechanistic relevance of these observations.

In this work, antibody functions measured in two cohorts of convalescent subjects were modeled using biophysical antibody profiles comprised of tandem attributes representing Fv-specificity and Fc characteristics. Multivariate linear regression identified distinct biophysical features that predicted antibody functions such as ADCC, ADCP, ADCD, and neutralization, showing the unique dependencies of each activity on different aspects of humoral responses. Although responses toward both endemic and pathogenic CoV were considered, models were almost exclusively reliant on SARS-CoV-2-specific responses in predicting functional activity. These predictions were robust and generalizable, performing similarly well in training and testing data subsets across cross-validation runs as in an independent validation cohort. The consistency between antibody features contributing to each modeled function and expected biological relevance suggests that modeling approaches such as that employed here can identify mechanisms of antibody activity, as has been observed in other studies^56-58^.

Spike-specific FcγR-binding antibodies made frequent contributions to models of effector functions, with FcγRIIa contributing strongly to phagocytosis and FcγRIIIa contributing strongly to NK cell activity. Among subclasses, IgG3 made an outsized contribution, consistent with prior studies in the context of other infections^59-61^, and monoclonal antibody subclass-switching studies^47,48^. In contrast, virus-specific IgM contributed to predictions of neutralization activity. SARS-CoV-2 specific IgM has attracted interest because of its association with lower risk of death from COVID-19^24^. Consistent with our experimental results, another study in which IgM was selectively depleted also observed resulting reduction in neutralization activity, but additionally confirmed the activity of the isolated IgM fraction^62,63^. Interestingly, SARS-CoV-2 specific IgM administered intranasally has been shown to be effective in treating novel SARS-CoV-2 variants of concern, including the alpha, beta, and gamma variants in a mouse model^64^. The finding that so much of the neutralizing activity of convalescent plasma against SARS-CoV-2 resides in the IgM fraction raises concern about that gamma globulin preparations may lose much of their antiviral activity as this isotype is removed. Similarly, the faster clearance profile of IgM as compared to IgG may hold implications for both frequency of dosing and timing of plasma donation.

While features contributing to functional predictions have both prior support from other studies and experimental validation within this cohort, other feature sets are likely to provide similar performance. Given high feature dimensionality and relatively fewer subjects, regularization was used to increase the quality of prediction. This approach simplified the resulting models, resulting in improved interpretability of the selected variables at the cost of eliminating features that are highly correlated to selected variables in the established model. Collectively, this modeling choice can result in a trade-off between model simplification and obscuring potential biological mechanisms. Other limitations include the use of surrogate functional assays that bear advantages in terms of throughput and reproducibility but pose limitations in terms of their biological relevance. As further functional assays reliant on free virions and infected cells are developed, it will be of interest to compare and contrast both the degree of correlation with these convenient proxy assays as well as to model those activities in pursuit of insights into unique subpopulations of antibodies that may be responsible for their induction, or to define general characteristics of a response that is highly polyfunctional.

As viral variants continue to emerge, rapid binding profiling may be an important complement to functional breadth assessments. Insights into how Fc characteristics of cross-reactive responses relate to diverse functions may provide accelerated insights into population-level susceptibility and support prioritization among candidate vaccine regimens. Numerous randomized clinical trials of convalescent plasma for COVID-19 are in the process of completion and it is likely that plasma remnants will be available for retrospective detailed serological analysis and correlation with clinical outcome^15^. This multivariate analysis provides a blueprint for carrying out such investigation, which could provide information on the antibody functions that contribute to clinical efficacy. The discovery of antibody functions associated with passive antibody efficacy could allow optimization of serological characteristics of mAbs, plasma and gamma globulin products for prevention and therapy of COVID-19.

## Materials and Methods

### Human subjects

The discovery cohort comprised 126 adult eligible convalescent plasma donors diagnosed with SARS-CoV-2 infection by nucleic acid amplification in the Baltimore, MD and Washington DC area (Johns Hopkins Medical Institutions, JHMI cohort) and has been previously described^27^. The validation cohort comprised 20 SARS-CoV-2 convalescent individuals from the Hanover, New Hampshire area (Dartmouth Hitchcock Medical Center, DHMC cohort)^43^. Infection with SARS-CoV-2 was confirmed in all convalescent subjects by nasopharyngeal swab PCR. Plasma (JHMI) or serum (DHMC) was collected from each donor approximately one month after symptom onset or first positive PCR test in the case of mild or asymptomatic disease. Samples from 15 naïve subjects collected from the Hanover, New Hampshire area served as negative controls. **Supplemental Table 1** provides basic clinical and demographic information for each cohort.

Human subject research was approved by both the Johns Hopkins University School of Medicine’s Institutional Review Board and the Dartmouth-Hitchcock Medical Center Committee for the Protection of Human Subjects. All participants provided written informed consent.

### Antibody features and functions

The magnitude, Fv specificity, and Fc domain characteristics of antibody responses to diverse coronavirus and control antigens were profiled by multiplexed Fc Array assay^22^, as previously described^26,43,65^. **Supplemental Table 2** reports the complete list of antigen specificities and Fc domain characteristics that were assayed. Fc Array data reported in median fluorescent intensity (MFI) was log transformed prior to analysis.

Antibody functions were assayed as previously described^26,43^. Briefly, neutralization of authentic virus^27,66,67^ was determined for samples from the JHMI cohort, whereas a pseudovirus neutralization assay^68^ was employed for evaluation of the DHMC cohort. Phagocytic activity was defined as the level of uptake of antigen-conjugated beads by THP-1 monocytes (ADCP)^69,70^ or primary neutrophils (ADNP)^71^. ADCC activity was modeled using a reporter cell line that expresses luciferase in response to FcγRIIIa ligation^72^. Antibody-dependent complement deposition was assessed by measuring C3b levels on antigen-conjugated beads following incubation in complement serum ^73^. For each assay, SARS-CoV-2 naïve samples were employed as negative controls, and data was collected in replicate.

### IgM depletion

IgM was depleted from serum as described previously^62^. Briefly, 200 μL of NHS HP SpinTrap resin (Cytiva) was equilibrated and used to immobilize anti-human IgM (μ-chain specific, Sigma I0759) at 850 μg/mL for 30 minutes with end-over-end mixing at room temperature. The resin was washed, quenched with 50 mM Tris HCl, 1M NaCl pH 8.0 and 0.1M sodium acetate 0.5 M NaCl pH 4, and incubated with serum diluted 1:5 in DMEM and incubated overnight at 4°C with end-over-end mixing. Flow-through was subsequently collected by centrifugation. IgG, IgA, and IgM levels of each selected sample were evaluated with and without IgM depletion by multiplex assay as described above^26,43,74^. Neutralization was measured by pseudovirus reporter assay as described above^68^.

### Data analysis and visualization

Basic analysis and visualization were performed using GraphPad Prism. Heatmaps, correlation plots, and other graphs were generated in R (supported by R packages pheatmap^75^, igraph^76^, and ggplot2^77^). Fc Array features were filtered by elimination of features for which the samples exhibited signal within 10 standard deviations (SD) of the technical blank. Log transformed SARS-CoV-2-related Fc Array features and selected functions were scaled and centered by their standard deviation from the mean (z-score) per cohort and visualized following hierarchical clustering according to Manhattan distance. A weighted correlation network of pairs of SARS-CoV-2-related features and selected functions for which Pearson’s correlation coefficient ≥0.5 was graphed.

Multivariate linear regression was employed to predict antibody functions based on biophysical features with the R package “Glmnet”^78^, as previously described^56-58^. Regularization by L1-penalization (LASSO) was applied to eliminate variables that were less relevant to the outcome by imposing a penalty on the absolute value of the feature coefficient in order to reduce overfitting and reinforce performance generalizibility^79^. Functional measurements of ADCP, neutralization, and S1-specific ADCD were log_10_ transformed to reduce the prediction error of the models based on the assumption that better fitting models were more likely to rely on biologically relevant features. The lambda parameter (λ) was tuned using five-fold cross-validation to minimize mean squared error. A process of 200-times repeated modeling was used to investigate the potential of the different combinations of the biophysical features for modeling. Established with the JHMI cohort, a final model was selected based on the median MSE obtained among the repeated run in the JHMI cohort. The selected features and their coefficients were reported at a value of λ at which median model performance fell one standard error above the minimum to optimize the generalizability and provide more regularization to the model. In the permutation test procedure, the penalized multivariate regression was performed against randomized functional outcomes in the JHMI cohort in a 200-time repeated fashion. The correlation network was conducted with the biophysical features that were repeatedly selected within the repeated modeling process.

## Data Availability

Data and code to reproduce analyses are available at (link pending).

## Data and Code Availability

Data and code to reproduce analyses are available at (link pending).

## Acknowledgements

The authors would like to thank the participants who generously agreed to contribute to this study, as well as the full study team. This work was supported in part by the Division of Intramural Research, NIAID, NIH.

## Author Contributions

Contributed samples – E.M.B., R.S., O.A., A.A.R.T., D.S., S.S., P.F.W.

Collected experimental data – H.N., A.R.C., S.E.B., K.L, S.E.B., R.S., O.A., W.W.-A., A.P., R.I.C.

Performed data analysis – H.N., S.X., R.I.C. Drafted the manuscript - H.N., S.X.

Reviewed and edited the manuscript – all authors

Supervised research – M.E.A., P.F.W. A.D.R., A.A.R.T., A.C., A.P.

Conceived of work – M.E.A.

## Conflict of Interest

The authors declare that they have no conflict of interest.

## Supplemental Figures and Tables

**Supplemental Figure 1.**
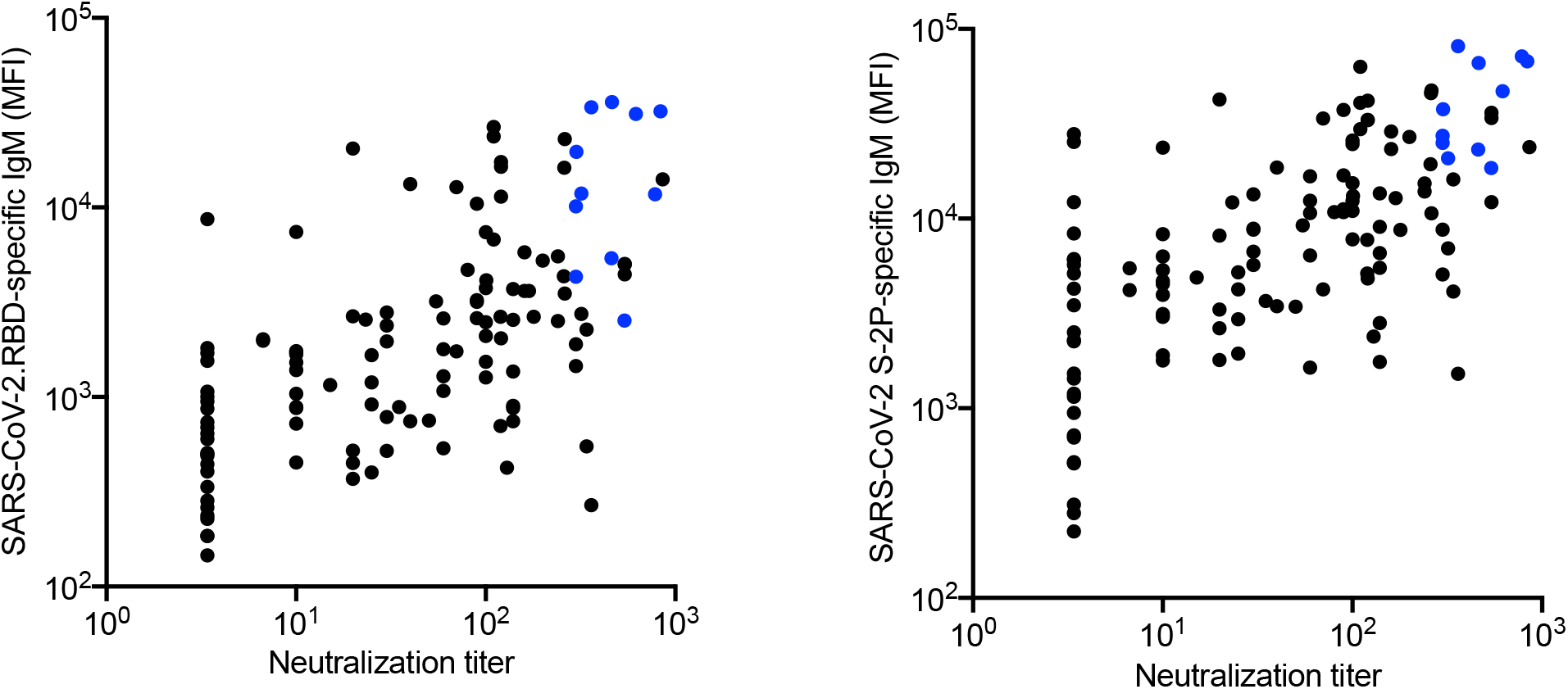
Samples selected for IgM depletion. Comparison of SARS-CoV-2 RBD-specific (left) and stabilized spike (S-2P)-specific IgM levels (median fluorescent intensity, MFI) and neutralization titer. Samples with high IgM and high neutralization titers that were selected for depletion are highlighted in blue.

**Supplemental Table 1.** Cohort characteristics. Demographic information on convalescent serum (DHMC) and convalescent plasma samples (JHMI). NA indicates not applicable and IQR indicates interquartile range. Reproduced from Natarajan, et. al, 2021^26^.

| Characteristic | JHMI Convalescent<br>n=126 | DHMC Convalescent<br>n=20 | Naive<br>n=15 |
| --- | --- | --- | --- |
| Median age (IQR), years | 42 (29-53) | 54 (45-62) | 34 (28-52) |
| Sex |  |  |  |
| Female | 58 (46%) | 10 (50%) | 8 (53.3%) |
| Male | 68 (54%) | 10 (50%) | 7 (46.7%) |
| Hospitalized (severity) |  |  |  |
| No | 114 (90.5%) | 16 (80%) | NA |
| Yes | 12 (9.5%) | 4 (20%) | NA |
| Median days since PCR+ or symptom onset (IQR) | 43 (38-48) | 38 (33-45) | NA |

**Supplemental Table 2.** Fc detection and antigen reagents.

| **Antigen** | **Source** | **Fc Detection** | **Source** |
| --- | --- | --- | --- |
| H1N1 HA1 | Immune Technology<br>IT-003-00110p | a- IgG | Southern Biotech 1030-09 |
| HSV gE | Immune Technology<br>IT-005-005p | a-IgG1 | Southern Biotech 9054-09 |
| SARS CoV-2 N | Immune Technology<br>IT-002-033Ep | a-IgG2 | Southern Biotech 9070-09 |
| SARS CoV-2 FP | New England<br>Peptide | a-IgG3 | Southern Biotech 9210-09 |
| SARS CoV-2 S1 (for Fc Array) | ACRO Biosystems<br>S1N-C52H3-100ug | a-IgG4 | Southern Biotech 9200-09 |
| SARS CoV-2 S1 (for ADCD) | Sino Biological<br>40150-V08B1-20 | a-IgA | Southern Biotech 2050-09 |
| SARS CoV-2 RBD | BEI Resources<br>NR-52366 | a-IgA1 | Southern Biotech 9130-09 |
| SARS CoV-2 S2 | Immune Technology<br>IT-002-034p | a-IgA2 | Southern Biotech 9140-09 |
| SARS CoV-2 S-2P | Expressed in Expi<br>293 | a-IgM | Southern Biotech 9020-09 |
| WIV1 S-2P | Expressed in Expi<br>293 | a-IgD | Southern Biotech <u>9030-09</u> |
| MERS S-2P | Expressed in Expi<br>293 | FcγR1 | Duke Protein Production Facility |
| MERS S1 | Sino Biological<br>40069-V08B1 | FcγR2a | Boesch, et. al, 2014 |
| HCoV OC43 S | Sino Biological<br>40607-V08B | FcγR2b | Boesch, et. al, 2014 |
| OC43 S-2P | Expressed in HEK<br>293F | FcγR3a | Boesch, et. al, 2014 |
| 229E S1 | Sino Biological<br>40605-V08H | FcγR3b | Boesch, et. al, 2014 |
| 229E S-2P | Expressed in Expi<br>293 |  |  |
| HKU1 S1 | Sino Biological<br>40606-V08H |  |  |
| HKU S-2P | Expressed in Expi<br>293 |  |  |
| NL63 S1 | Sino Biological<br>40604-V08H |  |  |

## Notes

### Competing Interest Statement

The authors have declared no competing interest.

### Author Declarations

Human subject research was approved by both the Johns Hopkins University School of Medicine Institutional Review Board and the Dartmouth-Hitchcock Medical Center Committee for the Protection of Human Subjects. All participants provided written informed consent.

## References

1 COVID-19 Vaccines, <https://www.fda.gov/emergency-preparedness-and-response/coronavirus-disease-2019-covid-19/covid-19-vaccines#eua-vaccines> (2021).

2 Iwasaki, A. What reinfections mean for COVID-19. The Lancet Infectious Diseases 21, 3–5, doi:10.1016/s1473-3099(20)30783-0 (2021).

3 Tillett, R. L. et al. Genomic evidence for reinfection with SARS-CoV-2: a case study. The Lancet Infectious Diseases 21, 52–58, doi:10.1016/s1473-3099(20)30764-7 (2021).

4 Lumley, S. F. et al. Antibody Status and Incidence of SARS-CoV-2 Infection in Health Care Workers. N Engl J Med 384, 533–540, doi:10.1056/NEJMoa2034545 (2021).

5 Harvey, R. A. et al. Association of SARS-CoV-2 Seropositive Antibody Test With Risk of Future Infection. JAMA Intern Med 181, 672–679, doi:10.1001/jamainternmed.2021.0366 (2021).

6 Letizia, A. G. et al. SARS-CoV-2 seropositivity and subsequent infection risk in healthy young adults: a prospective cohort study. The Lancet Respiratory Medicine, doi:10.1016/s2213-2600(21)00158-2 (2021).

7 Benner, S. E. et al. SARS-CoV-2 Antibody Avidity Responses in COVID-19 Patients and Convalescent Plasma Donors. J Infect Dis 222, 1974–1984, doi:10.1093/infdis/jiaa581 (2020).

8 Tobian, A. A. R. & Shaz, B. H. Earlier the better: convalescent plasma. Blood 136, 652–654, doi:10.1182/blood.2020007638 (2020).

9 Duan, K. et al. Effectiveness of convalescent plasma therapy in severe COVID-19 patients. Proc Natl Acad Sci U S A 117, 9490–9496, doi:10.1073/pnas.2004168117 (2020).

10 Shen, C. et al. Treatment of 5 Critically Ill Patients With COVID-19 With Convalescent Plasma. JAMA 323, 1582–1589, doi:10.1001/jama.2020.4783 (2020).

11 Libster, R. et al. Early High-Titer Plasma Therapy to Prevent Severe Covid-19 in Older Adults. N Engl J Med 384, 610–618, doi:10.1056/NEJMoa2033700 (2021).

12 O’Donnell, M. R. et al. A randomized double-blind controlled trial of convalescent plasma in adults with severe COVID-19. J Clin Invest 131, doi:10.1172/JCI150646 (2021).

13 Joyner, M. J. et al. Convalescent Plasma Antibody Levels and the Risk of Death from Covid-19. N Engl J Med 384, 1015–1027, doi:10.1056/NEJMoa2031893 (2021).

14 Klassen, S. A. et al. Convalescent Plasma Therapy for COVID-19: A Graphical Mosaic of the Worldwide Evidence. Front Med (Lausanne) 8, 684151, doi:10.3389/fmed.2021.684151 (2021).

15 Cohn, C. S. et al. COVID-19 convalescent plasma: Interim recommendations from the AABB. Transfusion 61, 1313–1323, doi:10.1111/trf.16328 (2021).

16 Group, R. C. Convalescent plasma in patients admitted to hospital with COVID-19 (RECOVERY): a randomised controlled, open-label, platform trial. Lancet 397, 2049–2059, doi:10.1016/S0140-6736(21)00897-7 (2021).

17 Simonovich, V. A. et al. A Randomized Trial of Convalescent Plasma in Covid-19 Severe Pneumonia. N Engl J Med 384, 619–629, doi:10.1056/NEJMoa2031304 (2021).

18 Agarwal, A. et al. Convalescent plasma in the management of moderate covid-19 in adults in India: open label phase II multicentre randomised controlled trial (PLACID Trial). BMJ 371, m3939, doi:10.1136/bmj.m3939 (2020).

19 Hung, I. F.-N. et al. Triple combination of interferon beta-1b, lopinavir–ritonavir, and ribavirin in the treatment of patients admitted to hospital with COVID-19: an open-label, randomised, phase 2 trial. The Lancet 395, 1695–1704, doi:10.1016/s0140-6736(20)31042-4 (2020).

20 Klingler, J. et al. Role of IgM and IgA Antibodies in the Neutralization of SARS-CoV-2. J Infect Dis, doi:10.1093/infdis/jiaa784 (2020).

21 Wang, Z. et al. mRNA vaccine-elicited antibodies to SARS-CoV-2 and circulating variants. Nature 592, 616–622, doi:10.1038/s41586-021-03324-6 (2021).

22 Widge, A. T. et al. Durability of Responses after SARS-CoV-2 mRNA-1273 Vaccination. N Engl J Med 384, 80–82, doi:10.1056/NEJMc2032195 (2021).

23 Edara, V. V. et al. Infection and Vaccine-Induced Neutralizing-Antibody Responses to the SARS-CoV-2 B.1.617 Variants. N Engl J Med, doi:10.1056/NEJMc2107799 (2021).

24 Atyeo, C. et al. Distinct Early Serological Signatures Track with SARS-CoV-2 Survival. Immunity 53, 524–532 e524, doi:10.1016/j.immuni.2020.07.020 (2020).

25 Guan, W. J. et al. Clinical Characteristics of Coronavirus Disease 2019 in China. N Engl J Med 382, 1708–1720, doi:10.1056/NEJMoa2002032 (2020).

26 Natarajan, H. et al. Markers of Polyfunctional SARS-CoV-2 Antibodies in Convalescent Plasma. mBio 12, doi:10.1128/mBio.00765-21 (2021).

27 Klein, S. et al. Sex, age, and hospitalization drive antibody responses in a COVID-19 convalescent plasma donor population. medRxiv, doi:10.1101/2020.06.26.20139063 (2020).

28 Tang, J. et al. Antibody affinity maturation and plasma IgA associate with clinical outcome in hospitalized COVID-19 patients. Nat Commun 12, 1221, doi:10.1038/s41467-021-21463-2 (2021).

29 Sterlin, D. et al. IgA dominates the early neutralizing antibody response to SARS-CoV-2. Sci Transl Med 13, doi:10.1126/scitranslmed.abd2223 (2021).

30 Liu, L. et al. Potent neutralizing antibodies against multiple epitopes on SARS-CoV-2 spike. Nature 584, 450–456, doi:10.1038/s41586-020-2571-7 (2020).

31 Toapanta, F. R. & Ross, T. M. Complement-mediated activation of the adaptive immune responses: role of C3d in linking the innate and adaptive immunity. Immunol Res 36, 197–210, doi:10.1385/IR:36:1:197 (2006).

32 Ward, E. S. & Ghetie, V. The effector functions of immunoglobulins: implications for therapy. Ther Immunol 2, 77–94 (1995).

33 Clark, M. R. IgG effector mechanisms. Chem Immunol 65, 88–110 (1997).

34 Atyeo, C. et al. Dissecting strategies to tune the therapeutic potential of SARS-CoV-2-specific monoclonal antibody CR3022. JCI Insight 6, doi:10.1172/jci.insight.143129 (2021).

35 Wec, A. Z. et al. Broad neutralization of SARS-related viruses by human monoclonal antibodies. Science 369, 731–736, doi:10.1126/science.abc7424 (2020).

36 Tortorici, M. A. et al. Ultrapotent human antibodies protect against SARS-CoV-2 challenge via multiple mechanisms. Science 370, 950–957, doi:10.1126/science.abe3354 (2020).

37 Suryadevara, N. et al. Neutralizing and protective human monoclonal antibodies recognizing the N-terminal domain of the SARS-CoV-2 spike protein. bioRxiv, doi:10.1101/2021.01.19.427324 (2021).

38 Ullah, I. et al. Live imaging of SARS-CoV-2 infection in mice reveals neutralizing antibodies require Fc function for optimal efficacy. bioRxiv, doi:10.1101/2021.03.22.436337 (2021).

39 Alter, G. et al. Collaboration between the Fab and Fc contribute to maximal protection against SARS-CoV-2 following NVX-CoV2373 subunit vaccine with Matrix-M vaccination. Res Sq, doi:10.21203/rs.3.rs-200342/v1 (2021).

40 Winkler, E. S. et al. Human neutralizing antibodies against SARS-CoV-2 require intact Fc effector functions for optimal therapeutic protection. Cell, doi:10.1016/j.cell.2021.02.026 (2021).

41 Adeniji, O. S. et al. COVID-19 Severity Is Associated with Differential Antibody Fc-Mediated Innate Immune Functions. mBio 12, doi:10.1128/mBio.00281-21 (2021).

42 Larsen, M. D. et al. Afucosylated IgG characterizes enveloped viral responses and correlates with COVID-19 severity. Science 371, doi:10.1126/science.abc8378 (2021).

43 Butler, S. E. et al. Distinct Features and Functions of Systemic and Mucosal Humoral Immunity Among SARS-CoV-2 Convalescent Individuals. Front Immunol 11, 618685, doi:10.3389/fimmu.2020.618685 (2020).

44 Wrapp, D. et al. Cryo-EM structure of the 2019-nCoV spike in the prefusion conformation. Science 367, 1260–1263, doi:10.1126/science.abb2507 (2020).

45 Alter, G. et al. High-resolution definition of humoral immune response correlates of effective immunity against HIV. Mol Syst Biol 14, e7881, doi:10.15252/msb.20177881 (2018).

46 Tomaras, G. D. et al. Vaccine-induced plasma IgA specific for the C1 region of the HIV-1 envelope blocks binding and effector function of IgG. Proc Natl Acad Sci U S A 110, 9019–9024, doi:10.1073/pnas.1301456110 (2013).

47 Chu, T. H. et al. Hinge length contributes to the phagocytic activity of HIV-specific IgG1 and IgG3 antibodies. PLoS Pathog 16, e1008083, doi:10.1371/journal.ppat.1008083 (2020).

48 Richardson, S. I. et al. IgG3 enhances neutralization potency and Fc effector function of an HIV V2-specific broadly neutralizing antibody. PLoS Pathog 15, e1008064, doi:10.1371/journal.ppat.1008064 (2019).

49 Heyman, B. S., Marc J. Structure, Function, and Production of Immunoglobulin M (IgM). Encyclopedia of Immunobiology 2, 1–14, doi:https://doi.org/10.1016/B978-0-12-374279-7.05001-3 (2016).

50 Fierz, W. & Walz, B. Antibody Dependent Enhancement Due to Original Antigenic Sin and the Development of SARS. Front Immunol 11, 1120, doi:10.3389/fimmu.2020.01120 (2020).

51 Henry, C., Palm, A. E., Krammer, F. & Wilson, P. C. From Original Antigenic Sin to the Universal Influenza Virus Vaccine. Trends Immunol 39, 70–79, doi:10.1016/j.it.2017.08.003 (2018).

52 Tauzin, A. et al. A single dose of the SARS-CoV-2 vaccine BNT162b2 elicits Fc-mediated antibody effector functions and T cell responses. Cell Host Microbe, doi:10.1016/j.chom.2021.06.001 (2021).

53 Holmes, M. J., Callow, K. A., Childs, R. A. & Tyrrell, D. A. Antibody dependent cellular cytotoxicity against coronavirus 229E-infected cells. Br J Exp Pathol 67, 581–586 (1986).

54 Cepica, A. & Derbyshire, J.B. Antibody-dependent cell-mediated cytotoxicity and spontaneous cell-mediated cytotoxicity against cells infected with porcine transmissible gastroenteritis virus. Can J Comp Med 47, 298–303 (1983).

55 Yilla, M. et al. SARS-coronavirus replication in human peripheral monocytes/macrophages. Virus Res 107, 93–101, doi:10.1016/j.virusres.2004.09.004 (2005).

56 Bradley, T. et al. Pentavalent HIV-1 vaccine protects against simian-human immunodeficiency virus challenge. Nat Commun 8, 15711, doi:10.1038/ncomms15711 (2017).

57 Pittala, S. et al. Antibody Fab-Fc properties outperform titer in predictive models of SIV vaccine-induced protection. Mol Syst Biol 15, e8747, doi:10.15252/msb.20188747 (2019).

58 Ackerman, M. E. et al. Route of immunization defines multiple mechanisms of vaccine-mediated protection against SIV. Nat Med 24, 1590–1598, doi:10.1038/s41591-018-0161-0 (2018).

59 Neidich, S. D. et al. Antibody Fc effector functions and IgG3 associate with decreased HIV-1 risk. J Clin Invest 129, 4838–4849, doi:10.1172/JCI126391 (2019).

60 Yates, N. L. et al. Vaccine-induced Env V1-V2 IgG3 correlates with lower HIV-1 infection risk and declines soon after vaccination. Sci Transl Med 6, 228ra239, doi:10.1126/scitranslmed.3007730 (2014).

61 Chu, T. H., Patz, E. F., Jr. & Ackerman, M. E. Coming together at the hinges: Therapeutic prospects of IgG3. MAbs 13, 1882028, doi:10.1080/19420862.2021.1882028 (2021).

62 Gasser, R. et al. Major role of IgM in the neutralizing activity of convalescent plasma against SARS-CoV-2. Cell Rep 34, 108790, doi:10.1016/j.celrep.2021.108790 (2021).

63 Klingler, J. et al. Role of Immunoglobulin M and A Antibodies in the Neutralization of Severe Acute Respiratory Syndrome Coronavirus 2. J Infect Dis 223, 957–970, doi:10.1093/infdis/jiaa784 (2021).

64 Ku, Z. et al. Nasal delivery of an IgM offers broad protection from SARS-CoV-2 variants. Nature, doi:10.1038/s41586-021-03673-2 (2021).

65 Morgenlander, W. R. et al. Antibody responses to endemic coronaviruses modulate COVID-19 convalescent plasma functionality. J Clin Invest 131, doi:10.1172/JCI146927 (2021).

66 Schaecher, S. R. et al. An immunosuppressed Syrian golden hamster model for SARS-CoV infection. Virology 380, 312–321, doi:10.1016/j.virol.2008.07.026 (2008).

67 Patel, E. U. et al. Comparative Performance of Five Commercially Available Serologic Assays To Detect Antibodies to SARS-CoV-2 and Identify Individuals with High Neutralizing Titers. J Clin Microbiol 59, doi:10.1128/JCM.02257-20 (2021).

68 Letko, M., Marzi, A. & Munster, V. Functional assessment of cell entry and receptor usage for SARS-CoV-2 and other lineage B betacoronaviruses. Nat Microbiol 5, 562–569, doi:10.1038/s41564-020-0688-y (2020).

69 Ackerman, M. E. et al. A robust, high-throughput assay to determine the phagocytic activity of clinical antibody samples. J Immunol Methods 366, 8–19, doi:10.1016/j.jim.2010.12.016 (2011).

70 McAndrew, E. G. et al. Determining the phagocytic activity of clinical antibody samples. J Vis Exp, e3588, doi:10.3791/3588 (2011).

71 Karsten, C. B. et al. A versatile high-throughput assay to characterize antibody-mediated neutrophil phagocytosis. J Immunol Methods 471, 46–56, doi:10.1016/j.jim.2019.05.006 (2019).

72 Gomez-Roman, V. R. et al. A simplified method for the rapid fluorometric assessment of antibody-dependent cell-mediated cytotoxicity. J Immunol Methods 308, 53–67, doi:10.1016/j.jim.2005.09.018 (2006).

73 Fischinger, S. et al. A high-throughput, bead-based, antigen-specific assay to assess the ability of antibodies to induce complement activation. J Immunol Methods 473, 112630, doi:10.1016/j.jim.2019.07.002 (2019).

74 Brown, E. P. et al. Multiplexed Fc array for evaluation of antigen-specific antibody effector profiles. J Immunol Methods 443, 33–44, doi:10.1016/j.jim.2017.01.010 (2017).

75 pheatmap: Pretty Heatmaps v. R package version 1.0.12 (2019).

76 Csardi GN. T. The igraph software package for complex network research. InterJournal Complex Systems, 1695 (2006).

77 Wickham, H. ggplot2: Elegant Graphics for Data Analysis. (Springer-Verlag, 2016).

78 Friedman J H. T.,, Tibshirani R. Regularization Paths for Generalized Linear Models via Coordinate Descent. Journal of Statistical Software 33, 1–22 (2010).

79 Tibshirani, R. Regression Shrinkage and Selection Via the Lasso. Journal of the Royal Statistical Society 73, 273–282 (2011).

